# Mapping geographic and demographic shifts in container breeding mosquito-borne disease transmission suitability in Central and South America in a warming world

**DOI:** 10.1101/2023.10.03.23296495

**Authors:** Sadie J. Ryan, Catherine A. Lippi, Anna M Stewart-Ibarra

## Abstract

The recent Intergovernmental Panel on Climate Change Sixth Assessment Report (IPCC-AR6) report brought into sharp relief the potential health impacts of a changing climate across large geographic regions. It also highlighted the gaps in available evidence to support detailed quantitative assessments of health impacts for many regions. In an increasingly urbanizing world, there is a need for additional information about the risk of mosquito-borne diseases from vectors adapted to human water storage behavior. Specifically, a better understanding of the geographic distribution of disease risk under different scenarios of climate warming and human populations shifts. For the Central and South America chapter of the IPCC Working Group II report, regional extractions of published projections of dengue and Zika risk in a changing climate were generated by one of the authors of this study. In that process, the lack of a compendium of available published risk estimates became apparent. This paper responds to that need and extends the scope of the IPCC report results for Central and South America. We present novel geospatial descriptions of risk for transmission for five mosquito-borne disease systems under future projected climate and demographic scenarios, including the potential risk for malaria in the event of the introduction and establishment of a vector of high global concern, *Anopheles stephensi*. We then present country-level and IPCC geospatial sub-region risk descriptions under baseline and future projected scenarios. By including demographic projections using the shared socioeconomic pathway (SSP) scenarios, we capture potential future risk in a way that is transparent and straightforward to compare and replicate. The goal of this paper is to report on these model output data and their availability. From a sub-regional perspective, the largest proportional gains in risk will be seen in the Southwestern South America (SWS) sub-region, comprising much of the southwestern coastline, for which suitability for *Aedes aegypti* transmitted dengue and Zika risk will see massive increases with warming, putting a large number of people at risk under future scenarios. In contrast, at the country level, the largest projected population risk impacts will be seen in Brazil for both arboviral and potential introduced malaria risk, despite some risks projected to decrease as parts of the country are too hot to sustain transmission risk. This paper provides modeled outputs for future use, in addition to broad summary descriptions at regional and country levels.

## Introduction

Ongoing and future climate change is expected to continue to have profound impacts on the global distribution and burden of many mosquito-borne diseases (MBDs) [1–5]. Nearly every link in the transmission chain of vector-borne pathogens is mediated by environmental influences, notably on vector reproduction and behavior, and pathogen replication [1,4]. The ideal conditions that drive vector-borne transmission systems vary with different vector-pathogen combinations. Still, many MBDs of public health importance, including malaria, yellow fever, dengue fever, and Zika, are typically associated with the tropics [1].

Increasing global temperatures are projected to expand the suitable transmission range of MBDs in some areas. However, these changes are not expected to result in uniformly elevated transmission risk throughout all regions. Rather, we expect that there will be shifts in the geographic expanse of MBD thermal transmission suitability [5]. As subtropical and temperate areas warm, becoming more suitable for the transmission of historically tropical diseases, other locations currently associated with high MBD incidence may become too hot, exceeding the physiological thermal limits of pathogens and vectors. While decreasing MBD suitability may seem advantageous at face value, any decrease in risk may be grossly outweighed by cumulative gains in the geographic area of suitability, the extension of transmission season length in locations that currently experience low MBD burden, or the expansion of the geographic range of transmission suitability into high population centers. It is also vital for public health planners to understand the shifting risk of MBDs against the backdrop of not only a changing climate, but also the movement of people on the landscape.

As the world experiences climate warming, we are simultaneously experiencing changes to the global human population, such as increasing urbanization, population densities, contact rates, and movements of people, goods, pathogens, and vectors. These changes directly affect MBD risk, particularly in urban and periurban environments. The spread of novel mosquito vectors has, in many instances, been linked to international trade networks and the movement of goods. For example, the expansion of *Ae. albopictus* via maritime transport, the used tire trade, and ground transportation networks [6], and recent detections of *An. stephensi* near seaports [7], and in a livestock quarantine station [8]. Human water storage practices and behaviors can create reservoirs of suitable habitat for mosquito oviposition on landscapes with otherwise inadequate precipitation. Water use and storage practices have consistently been identified as a major risk factor for urban dengue throughout Latin America, influencing household risk of infection [9–11], mosquito presence [12–17], and driving vector activity in arid cities [18]. Because of the profound influence that human activity has on vector distributions and MBD risk, it is therefore prudent to consider the intersection of thermal suitability and socioeconomic factors when estimating overall future vector suitability and population-level risk to humans.

Previous studies have made efforts to delineate potential shifts in the geography of MBD transmission risk driven by climate change [2,4,19,20]. Much of this work has focused on the transmission of *Plasmodium falciparum* malaria in Africa, given its considerable global health burden. Although malaria remains a public health concern in the Americas, outbreaks of arboviral diseases currently place larger resource strains on many public health systems throughout Latin America. Dengue fever is a major public health issue throughout Central and South America. In 2019, the Pan American Health Organization (PAHO) reported more than 3.1 million cases in the Americas, which included 28,176 severe cases and 1,535 deaths [21,22].

Following the unprecedented 2019 outbreak, dengue fever outbreaks continue to burden public health systems, which also had to contend with the devastating effects of the SARS-CoV-2 pandemic [22]. Zika virus, which swept through the region in 2016, imposed a high burden throughout Latin America as an emerging pathogen. While cases have dropped precipitously in many countries following the pandemic, Zika is still circulating in the Americas (over 40,000 cases reported in 2022 [23], and has the potential to trigger future outbreaks as population-level immunity wanes.

Malaria incidence has declined greatly throughout the Americas in recent decades, where cases have dropped by 70% between 2000 and 2021 [24]. Historically, *P. falciparum* accounted for the bulk of regional cases in Latin America, though following eradication efforts, most malaria burden is currently attributable to *P. vivax* [24,25]. Several regions in Central and South America have successfully achieved local malaria elimination through concerted vector control efforts, including Belize [26], El Salvador [27], Argentina, Paraguay [28], and the Ecuadorian- Peruvian border [29]. Nevertheless, localized hotspots of malaria activity persist, and have even increased, throughout the region. The reemergence of malaria remains a concern for public health officials [30], particularly in the Amazon region [25], which includes portions of southern Venezuela and Brazil [31,32].

In addition to the current threat of reemergence, the invasive mosquito *Anopheles stephensi* would further complicate the landscape of malaria transmission in the Americas should this vector become established. Global projections of thermal suitability already indicate that the Americas are suitable for establishment and transmission, should the vector be introduced [33]. Adept at transmitting both *P. falciparum* and *P. vivax*, *An. stephensi* has aggressively expanded its range in recent years and is associated with increased malaria outbreaks in the Horn of Africa [34]. Perhaps most troublesome, this vector has life history characteristics that mirror container-breeding *Aedes spp.*, presenting the unprecedented risk of urban malaria transmission in its introduced range [34].

Studies focusing on exposure risk solely by geographic region or landscape suitability may not communicate the most useful information for public health planning. Expressing risk in terms of estimated population exposure, or people at risk (PAR), clearly demonstrates to stakeholders the potential impact of shifting MBD transmission suitability on health infrastructure. This is a salient point for informing policy in Latin America, where public health authorities grapple with limited resources in the face of large MBD outbreaks. Further to this point, many of Latin America’s major population centers are located in coastal areas, and uneven population distributions mean that assessing potential exposure to risk purely in terms of geographic area does not capture the nuance of projected health burdens.

This study aims to generate model-based descriptions of container breeding MBD risk for IPCC- designated sub-regions in Central and South America, and country-level summary information under baseline and future climate projections and using corresponding population projections from the Global Population of the World (GPW4) and Shared Socioeconomic Pathways (SSPs) products. This work is the first effort to bring mapped temperature suitability models for currently circulating arboviral vector and disease risks and malaria transmission under the scenario of a potential novel vector (*An. stephensi*) together in one framework, facilitating comparisons of risk across diseases, climate change scenarios, and countries in these regions. In order to make these results available and useful for decision-making and planning, the resulting rasterized model outputs, and both IPPC sub-regional and country-level data are freely and openly available via the Harvard Dataverse (XXX URL upon publication).

## Materials and Methods

### Geographic coverage

In this study, we projected models of thermal suitability for MBDs to Central and South America, describing risk for two arboviral diseases with vectors that are well established in the Americas (i.e., dengue and Zika), in addition to two potential scenarios of malaria transmission by invasive *An. stephensi*, which is not currently present in Latin America. In addition to describing risk by country, we also summarized model output by geographic sub- regions, as defined by the Intergovernmental Panel on Climate Change (IPCC), which encompass 19 countries (Table 1, and seen in Figs 1,2,3,5,6). The 8 IPCC reference sub- regions used in this study are Southern Central America (SCA), Northern South America (NSA), Northwestern South America (NWS), Northeastern South America (NES), Southwestern South America (SWS), Southeastern South America (SES), Southern South America (SSA), and South American Monsoon (SAM) [35].

**Fig. 1.**
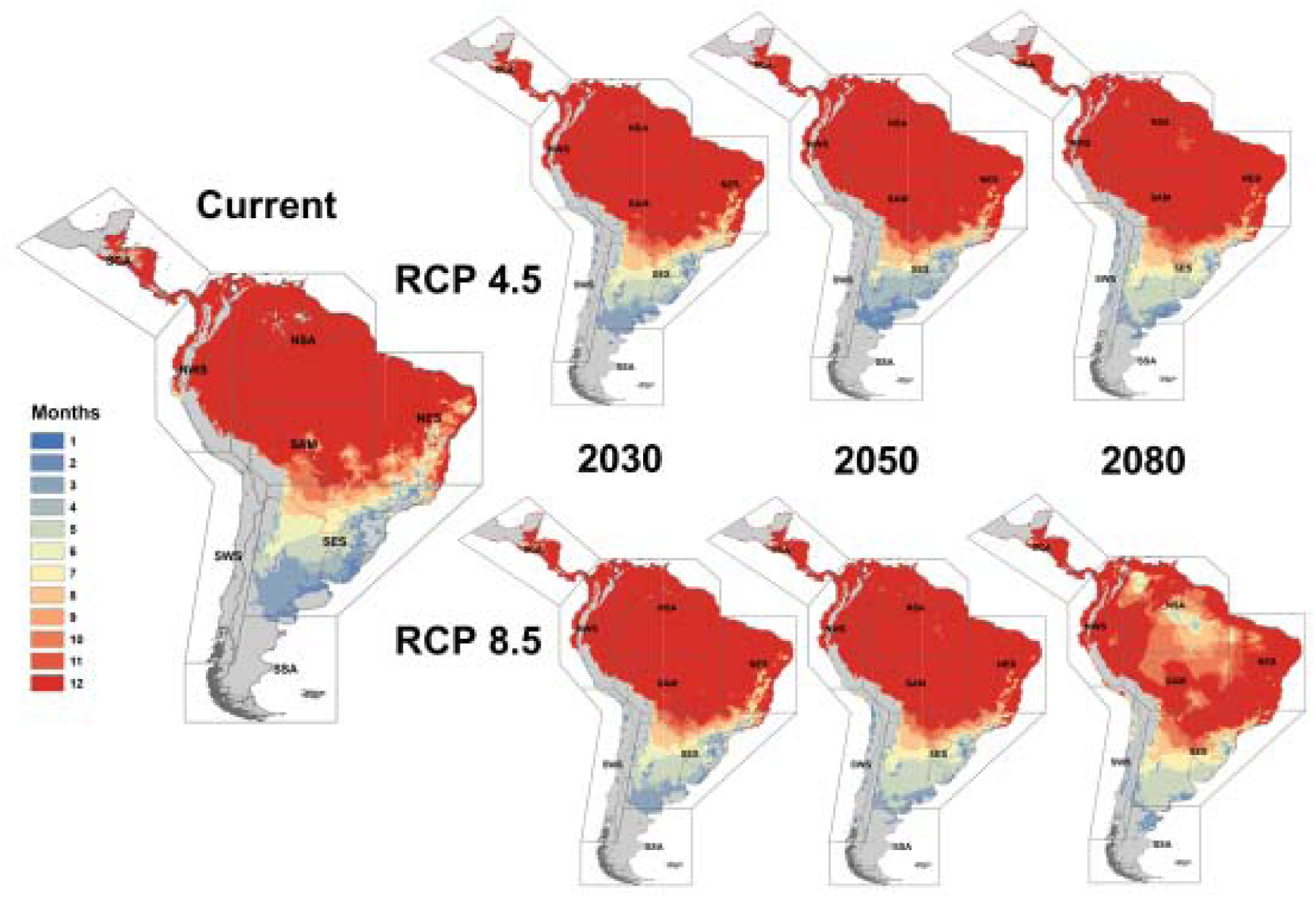
Thermal suitability for dengue transmitted by *Aedes aegypti*.

**Figure 2.**
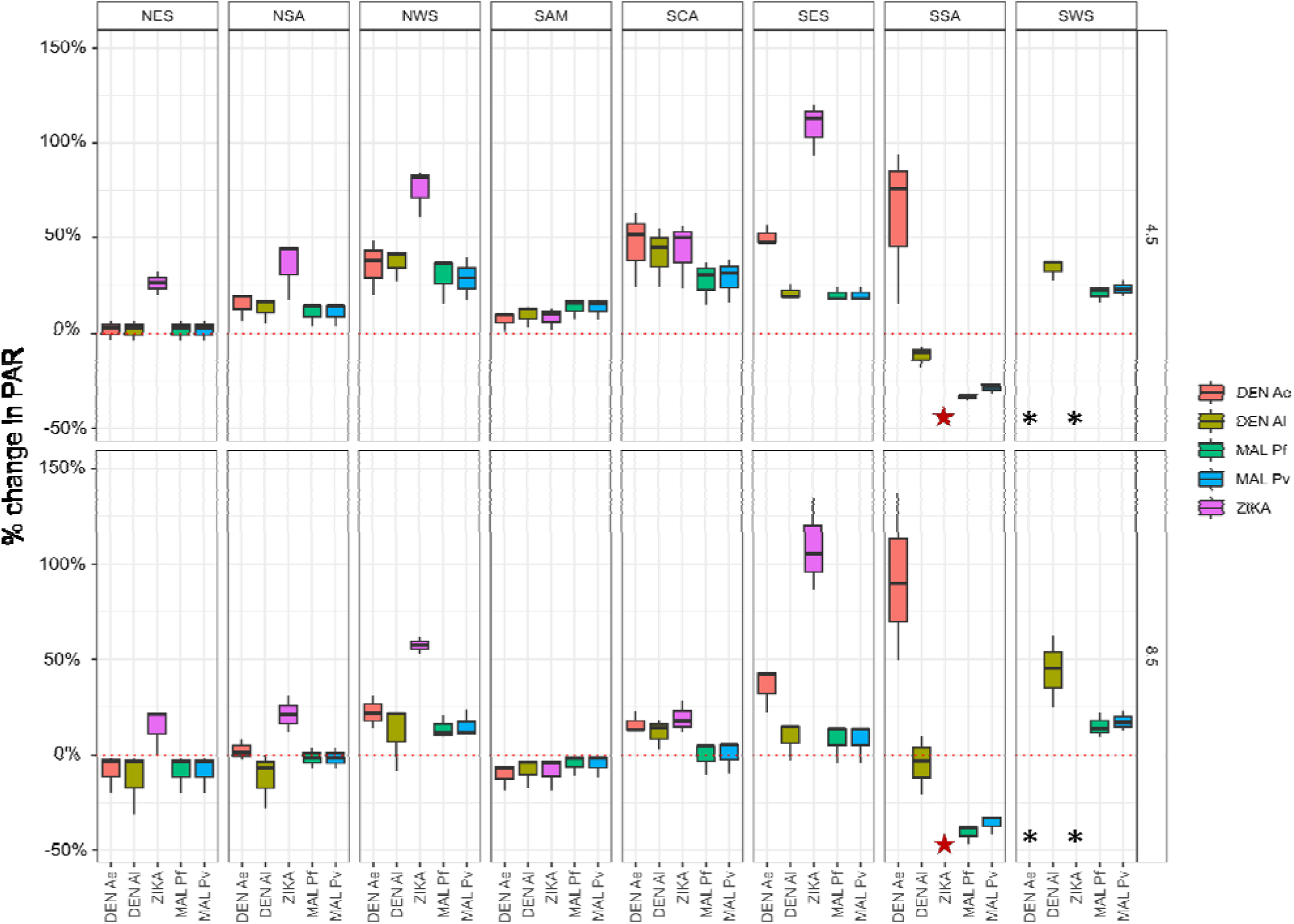
The percent of change in people at risk (PAR) from baseline to 2030, 2050, 2080 (given as boxplot) under the two climate projections, RCP 4.5 and RCP 8.5, for the five potential MBDs in this study, across the 8 sub-regions of Central and South America defined by IPCC for one or more months of suitability. DEN Ae = dengue transmitted by *Aedes aegypti*; DEN Al = Dengue transmitted by *Aedes albopctus;* MAL Pf = *Plasmodium falciparum* malaria transmitted by *Anopheles stephensi*; MAL Pv= *P. vivax* malaria transmitted by *An. stephensi*; ZIKA = Zika virus transmitted by *Ae. aegypti.* **Note**: at baseline there is no predicted risk for ZIKA in SSA, so all change is infinite (red star); SWS, changes exceeded 150% for DEN Ae and ZIKA (black asterisks).

**Table 1.** Central and South American countries with associated IPCC sub-regions.

| Region | Country | IPCC Sub-region |
| --- | --- | --- |
| Central America | Belize | SCA |
|  | Costa Rica | SCA |
|  | El Salvador | SCA |
|  | Guatemala | SCA |
|  | Honduras | SCA |
|  | Nicaragua | SCA |
|  | Panama | SCA, NWS |
| South America | Argentina | SES, SWS, SSA |
|  | Bolivia | SAM, SES, SWS |
|  | Brazil | NSA, NES, NWS, SAM, SES |
|  | Chile | SWS, SSA |
|  | Colombia | NWS |
|  | Ecuador | NWS |
|  | Guyana | NSA |
|  | Paraguay | SES, SAM |
|  | Peru | NWS, SAM, SWS |
|  | Suriname | NSA |
|  | Uruguay | SES |
|  | Venezuela | NSA, NWS |

### Thermal suitability models

We geographically projected published, temperature-driven mechanistic models to map the thermal suitability of mosquito-borne pathogen transmission for five vector-pathogen combinations. These included models of dengue virus transmission by *Aedes aegypti* and *Aedes albopictus* [36], Zika virus transmission by *Ae. aegypti* [37], and models of two malaria parasites (*Plasmodium falciparum* and *P. vivax*) transmitted by *An. stephensi* [38]. In this study, we used the critical thermal (temperature) bounds for dengue transmission by *Aedes aegypti* and *Ae. albopictus*, identified in Mordecai et al. 2017 [36], Zika transmission from Tesla et al., 2018 [37], and malaria transmission by *An. stephensi* from Villena et al. 2022 [38] to map thermal transmission suitability in Central and South America, taking values where the temperature-dependent transmission suitability metric for each mosquito:pathogen was greater than zero (S(T) > 0), with posterior probabilities greater than 0.975. The thermal limits used for dengue transmission are as follows: 21.3-34.0°C by *Ae. aegypti,* and 19.9 - 29.4°C by *Ae. albopictus*; for Zika transmitted by *Ae. aegypti*: 23.9 - 34.0°C; and for malaria transmitted by *An*. *stephensi*: 16.0 - 36.5°C for *P*. *falciparum,* and 16.6 - 31.7°C for *P*. *vivax*.

### Climate data

We used the WorldClim v1.4 product as our baseline climate data for model projections. Climate model output data for future scenarios were downloaded from the Consultative Group for International Agricultural Research (CGIAR) research program on Climate Change, Agriculture, and Food Security (CCAFS) web portal (http://ccafs-climate.org/data_spatial_downscaling/). All climate data were obtained at a resolution of 5-arc minutes. We used climate data inputs at three time horizons: 2030, 2050, and 2080, to project thermal models, using ensemble forecasts from the AR5 Archive under two representative concentration pathways (RCP 4.5 and RCP 8.5), and four general circulation models (GCMs) following methods described in Ryan et al. 2020 and Ryan, Lippi, and Zermoglio 2020 [19,39]. The GCMs used in this study were the Beijing Climate Center Climate System Model (BCC- CSM1.1); the Hadley GCM (HadGEM2-AO and HadGEM2-ES); and the National Center for Atmospheric Research’s Community Climate System Model (CCSM4).

*Population data* – We used the 2015 Global Population of the World (GPW) gridded population as our baseline for population descriptions. For the future population projections, we chose to use the Shared Socioeconomic Pathways (SSPs) projections [40–42] for the following plausible combinations: RCP 4.5 x SSP2 and RCP 8.5 x SSP5 [43]. We thus selected the corresponding projected populations for 2030, 2050, 2080 for each RCPxSSP combination.

*Mapping population at risk* – Risk maps incorporating climate information can be an important decision-support tool for the public health sector, part of an integrated climate adaptation strategy. To facilitate the utility of this type of geospatial modeling output for public health decision-making, and regional planning, it is important to generate information using the geographic designations from major decision-making bodies, but also provide country-level summary information.We summarized ‘risk’ in terms of the number of people subject to x months of transmission-suitable mean temperature, for dengue (both for *Ae. aegypti* and *Ae. albopictus* transmission) following Ryan et al. 2019 [2], for Zika transmission by *Ae. aegypti* following Ryan et al. 2021 [2,20], and for transmission of *P. falciparum* and *P. vivax* by *An. stephensi* following Ryan et al. 2023 [33]. We summarized PAR by both country and IPCC climate reference regions for Central and South America. For ease of interpretation, we followed methodology similar to that in Ryan et al. 2019, 2021, and 2023, summarizing the risk in terms of year-round transmission suitability (12 months), or any risk (one or more months of suitability) [2,20,33]. Mechanistic transmission models were projected onto climate data in R (v. 4.1.2) with the package ‘raster’ [44] (including some operations updated to use package ‘terra’ [45]), and monthly mean temperatures were thresholded according to the thermal suitability limits for each vector:pathogen pairing. The number of suitable months of transmission was then summed (0– 12) in a pixel-wise analysis for Central and South America, and monthly suitability maps were produced for baseline and future scenarios using ArcGIS ver. 10.1 [46].

## Results

### Dengue transmitted by Aedes aegypti

Under current climate conditions, much of the NSA, NWS, SAM, and NES regions are suitable for year-round dengue transmission by *Ae. aegypti* mosquitoes (Fig. 1). At baseline climate and population projections, there are collectively more than 350 million PAR of exposure to one or more months of thermal transmission suitability for dengue transmission by *Ae. aegypti* (Table 2). We found that the SES region, which spans Uruguay and portions of southern Brazil, Bolivia, Paraguay, and Argentina, has the highest PAR, with over 120.5 million. Under all realizations of climate and population scenarios, this risk increase ranges from 22-57% (Fig. 2).

**Table 2.**
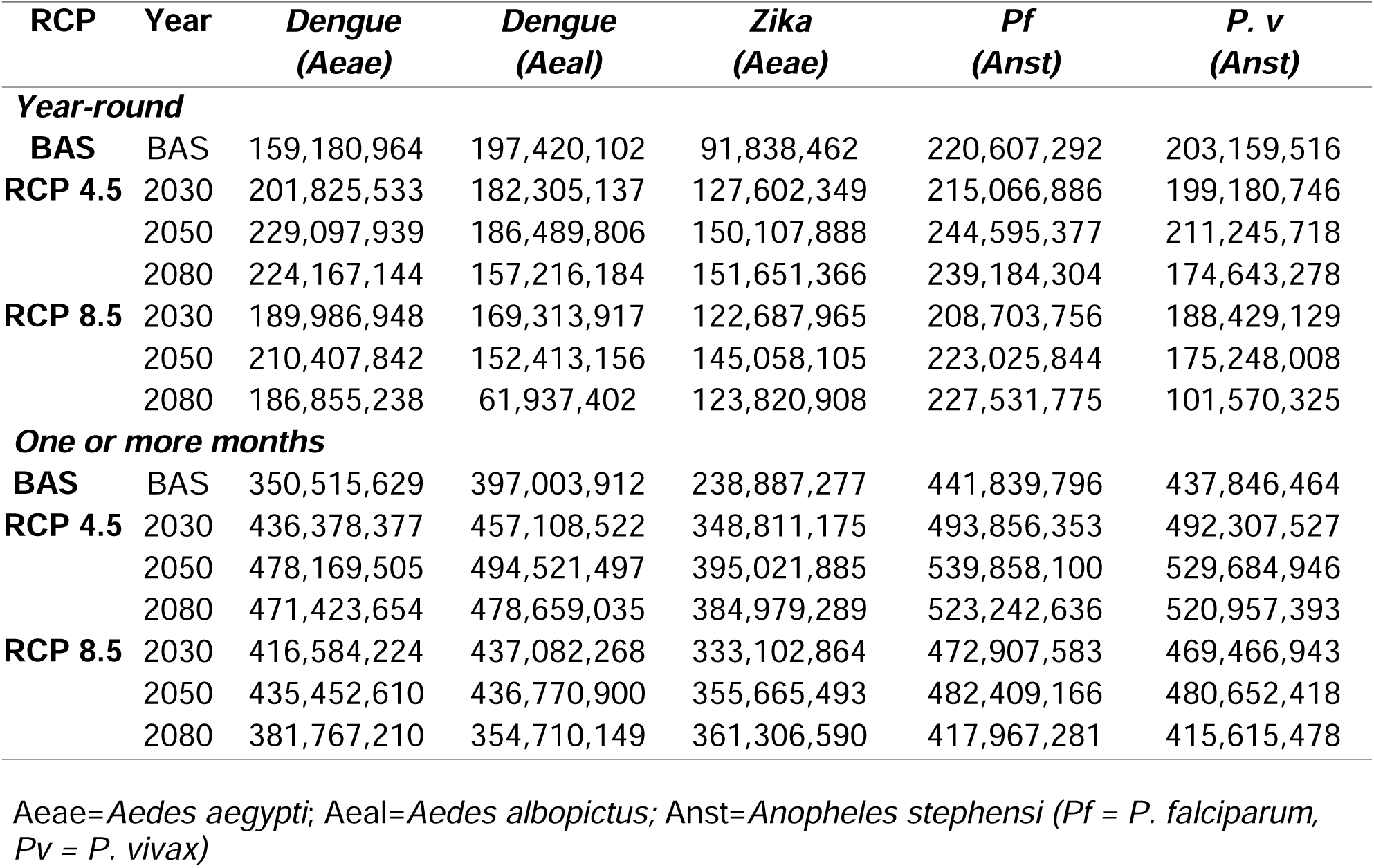
Total People at Risk (PAR) for thermal suitability for transmission (i.e. dengue virus transmitted by *Ae. aegypti* or *Ae. albopictus*, and Zika virus transmitted by *Ae. aegypti*) in Central and South America, and potential (PAR) for thermal suitability of malaria transmission (i.e. *P. falciparum* or *P. vivax* transmitted by *An. stephensi*), at baseline climate (BAS), and under two representative concentration pathways (RCP 4.5 and RCP 8.5) paired with a shared socioeconomic pathway projection of population growth (RCP 4.5LJ×LJSSP2; RCP 8.5LJ×LJSSP5).

In contrast, the SWS region, spanning much of Chile and the Western coastal region of South America, has the second lowest baseline population risk at around 2.4 million people. Yet, the SWS region has the highest project increases in PAR, ranging from increases of 204-374% (Fig. 2), as changing climate and population distributions coincide.

Conversely, the NES region, comprising northeastern Brazil, has a high baseline of 86.6 million PAR for one or more months of transmission. The NES region is largely characterized by up to 12 months of suitability under current climate conditions, with 54.5 million PAR of year-round transmission. Future changes in PAR for the NES region range from small increases in PAR (6%) to declining risk (-20%) (Fig. 2), due to a combination of higher temperatures and population shifts in response to changing climate.

Examining year-round suitability by country, Brazil has the highest risk of exposure under baseline conditions with over 60 million PAR, which markedly increases under future model projections (Fig. 5A). Colombia, Venezuela, and Ecuador are projected to experience similar patterns of increasing PAR, though increasing from much lower baseline populations.

### Dengue transmitted by Aedes albopictus

Under current climate conditions, much of the NSA, NWS, SAM, and NES regions are suitable for year-round dengue transmission by *Ae. albopictus* (Fig. 3). Collectively, there are more PAR of dengue transmission by *Ae. albopictus* than by *Ae. aegypti*, with over 397 million PAR for one or more months of transmission at baseline (Table 2). This total notably includes Brazil, the country with the highest at-risk population under baseline climate conditions, with over 190 million PAR for any level of transmission, and over 83 million PAR for year-round transmission (Fig. 5B).

**Fig. 3.**
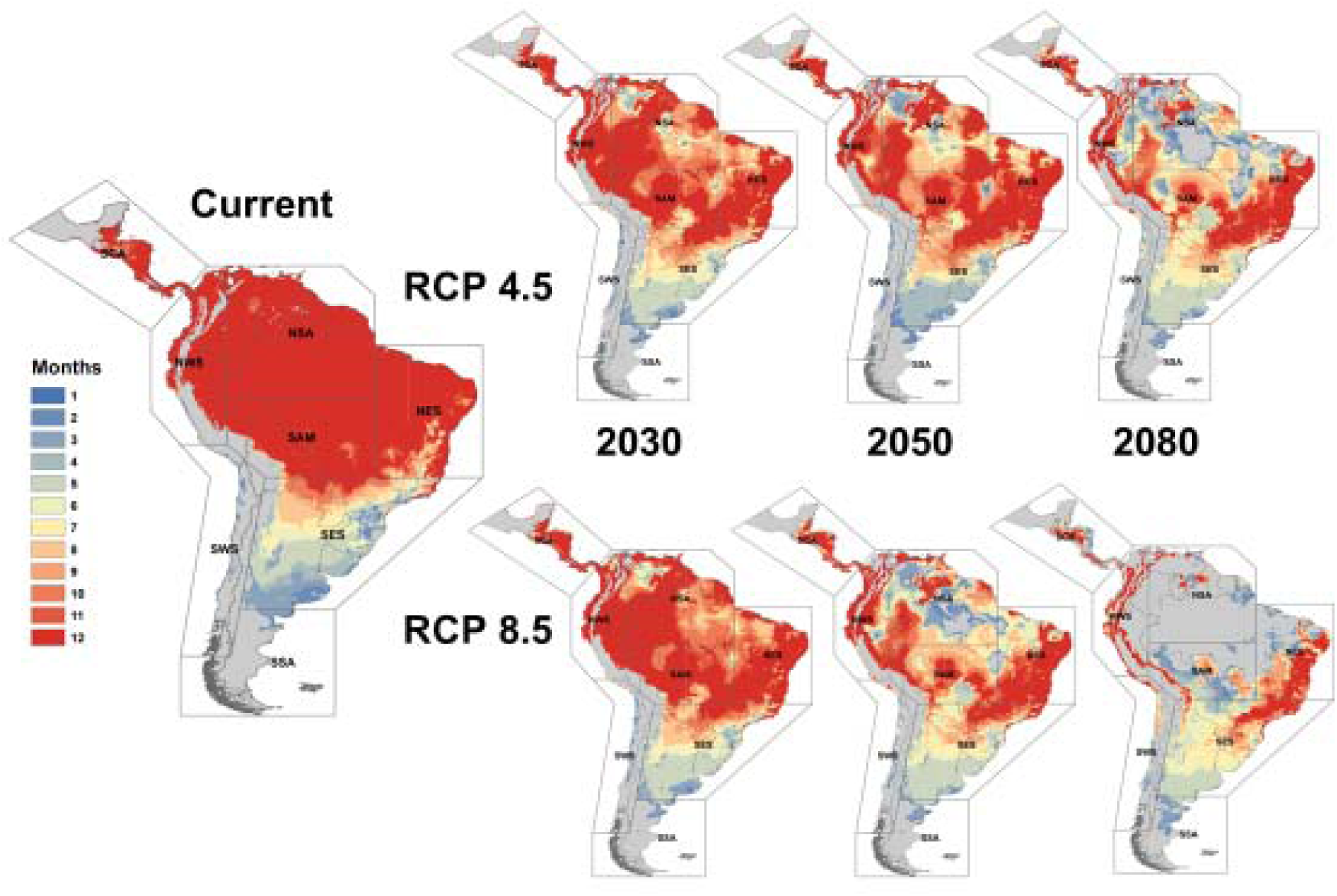
Thermal suitability for dengue transmitted by *Aedes albopictus*.

In contrast with dengue transmitted by *Ae. aegypti*, areas of thermal suitability for year-round *Ae. albopictus* transmission are projected to decrease markedly throughout the NSA, NWS, NES, SAM, and SES regions. Some of these reductions are also reflected in PAR projections for one or more months of transmission, where the biggest decreases in transmission exposure were -20.6 (SSA region under RCP 8.5), -27.9% (NSA region under RCP 8.5), and -31.4% (NES region under RCP 8.5) (Fig. 2). These range contractions coincide with warming temperatures, which will exceed the thermal limits for transmission by *Ae. albopictus* throughout much of these regions in the future.

Despite reductions in year-round PAR, projections for any dengue transmission (i.e., one or more months) by *Ae. albopictus* are projected to increase in many scenarios, by as much as 41.9% (NWS region under RCP 8.5) to 54.5% (SCA region under RCP 4.5) (Fig. 2). Notably, much of the SCA region in Central America, and coastal portions of the NWS and NES regions will remain suitable for year-round transmission of dengue, regardless of the scenario, where countries including Ecuador and Guatemala will experience slight increases in PAR (Fig. 5B).

### Zika transmitted by Aedes aegypti

Transmission of Zika by *Ae. aegypti* shows different spatial patterns of thermal suitability compared to dengue transmission. Under current climate conditions, year-round transmission of Zika is possible throughout most of the NSA region (Fig. 4). The NWS, NES, SCA, and SAM regions also have distinct areas at risk of year-round Zika transmission, however, these areas are much reduced compared to present-day dengue risk. Likewise, baseline PAR for Zika transmission is lower than that of dengue, ranging from approximately 91.8 million PAR for year- round Zika transmission, and over 238 million PAR for any Zika transmission (Table 2).

**Fig. 4.**
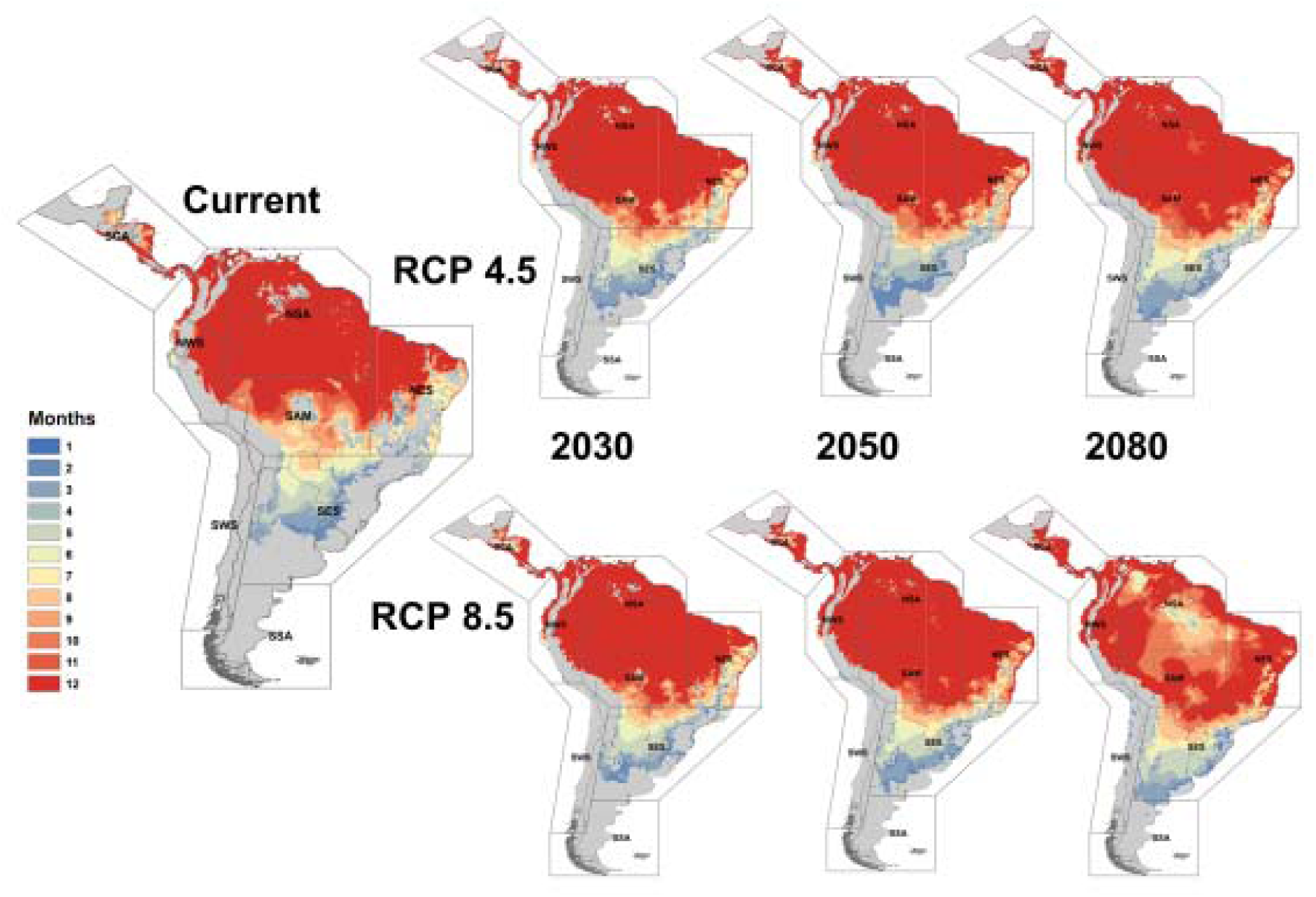
Thermal suitability for Zika transmitted by *Aedes aegypti*.

However, there are considerable gains in areas that will become suitable for Zika transmission in the future under nearly every scenario of climate change, and this is reflected in collective net increases in PAR (Table 2). Though baseline PAR is considerably lower for Zika transmission than dengue, the number of people exposed to potential transmission in the future will increase across the five countries with the highest PAR (Fig. 5C). Under future conditions, large portions of the NES, NWS, SAM, and SCA regions will see increased transmission suitability, and this will correspond with increasing PAR in most scenarios (Fig. 2). A long-term estimate under the most extreme scenario of climate change (i.e., 2080 under RCP 8.5) predicts shifting risk in NSA and SAM regions, where there are some reductions in estimated suitability, though overall suitability will still be greater than present-day conditions.

**Fig. 5.**
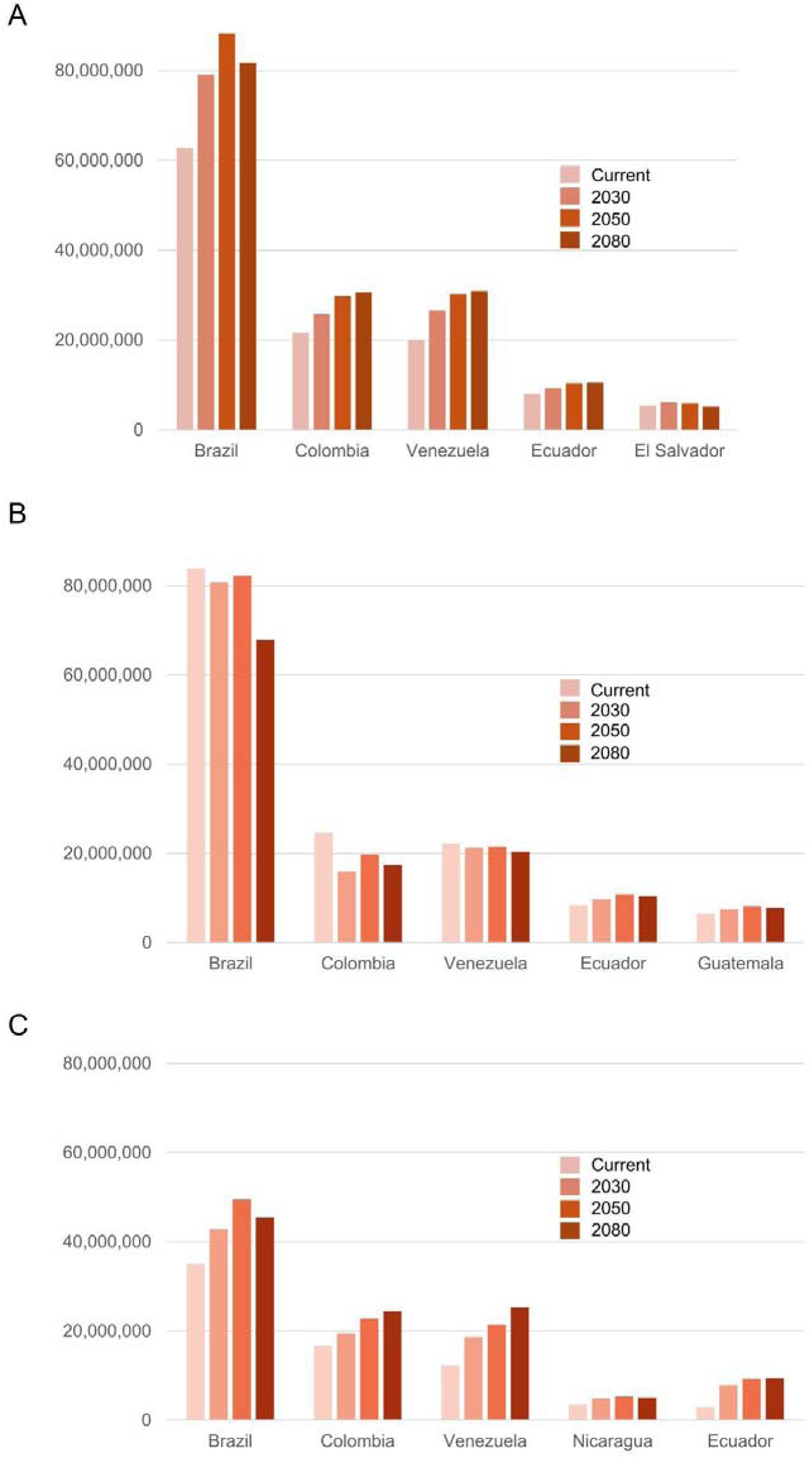
Top five countries in Central and South America with the highest population at risk (PAR) for exposure to year-round dengue transmitted by *Ae. aegypti* (A), dengue transmitted by *Ae. albopictus* (B) and Zika transmitted by *Ae. aegypti.* PAR will change in the future as geographic suitability shifts under climate change (RCP 4.5 shown).

### Plasmodium falciparum transmitted by Anopheles stephensi

Many regions in Central and South America will be suitable for transmission of *P. falciparum* by *An. stephensi*, both now and in the future if the vector invades and becomes established in the region (Fig. 6). The NSA, NWA, SAM, and NES regions are almost entirely suitable for year-round transmission risk, as are northern and central portions of the SES region spanning parts of Brazil, Paraguay, and Argentina. Notably, areas of suitable transmission extend further south compared to those of the arboviruses, where the length of the transmission season for *P. falciparum* malaria will become extended throughout much of Argentina. Nearly all of the SCA region is suitable for year-round *P. falciparum* transmission, putting more of this area at risk compared to arboviruses. Baseline totals for PAR of transmission exceed the totals for arboviruses, with over 220 million PAR of year-round transmission and over 441 million PAR for one or more months (Table 2). Under climate projections, shifting suitability is projected to initially result in decreased PAR for transmission in some regions in the near future, particularly under RCP 8.5, with decreases of up to -37.6% in the SSA region by 2030 (Fig. 2). However, by 2050 PAR could increase in every region under RCP 4.5, with the exception of SSA, with gains in PAR ranging from 5.8% (NES) to 36.6% (NWS) (Fig. 2). Brazil, the country with the highest baseline PAR, is projected to have modest declines in risk, while other countries including Venezuela and Colombia, will see marked increases in their at-risk population (Fig. 8A).

**Fig. 6.**
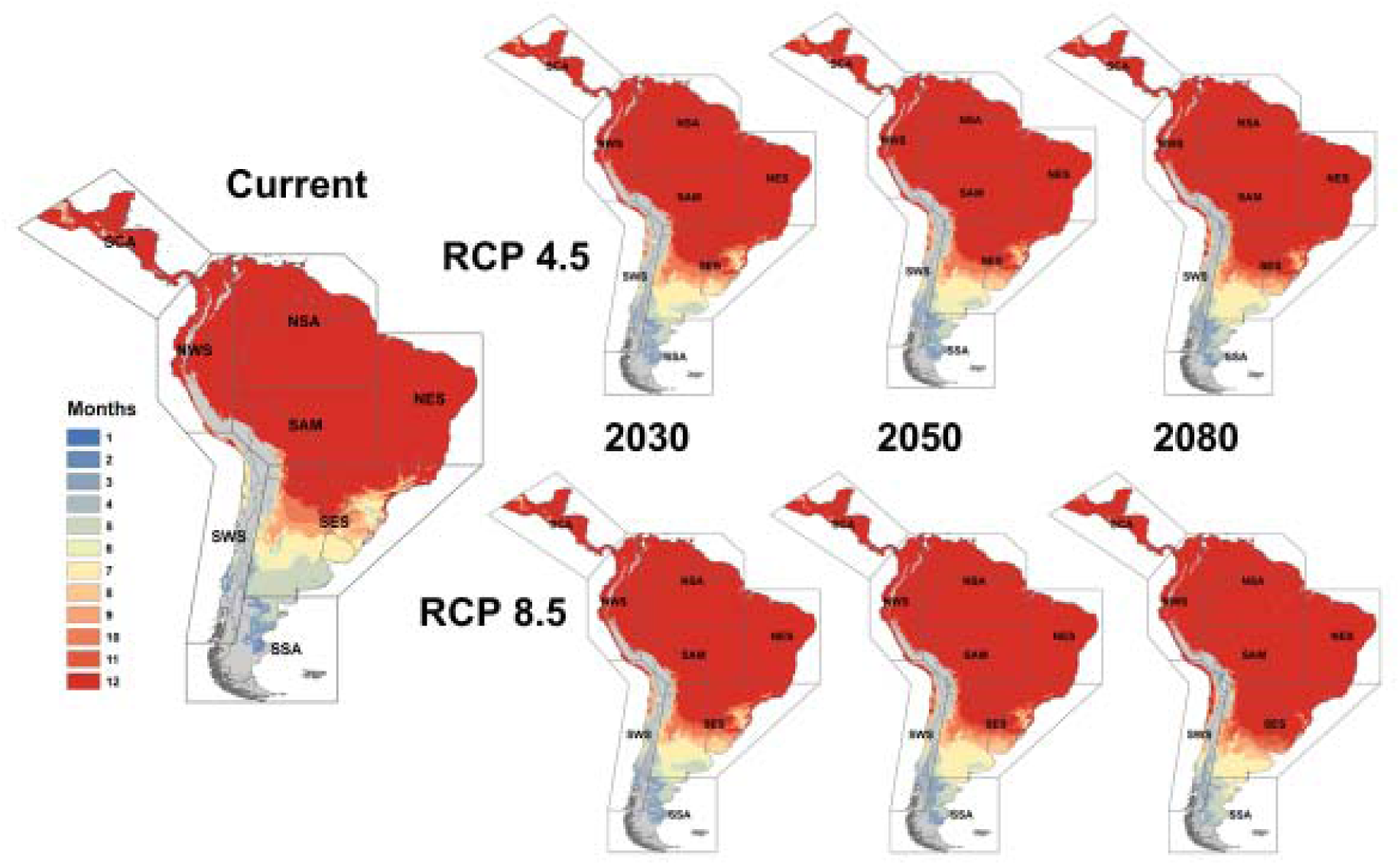
Thermal suitability for transmission of *Plasmodium falciparum* malaria by the potential invasive *Anopheles stephensi*.

**Fig. 7.**
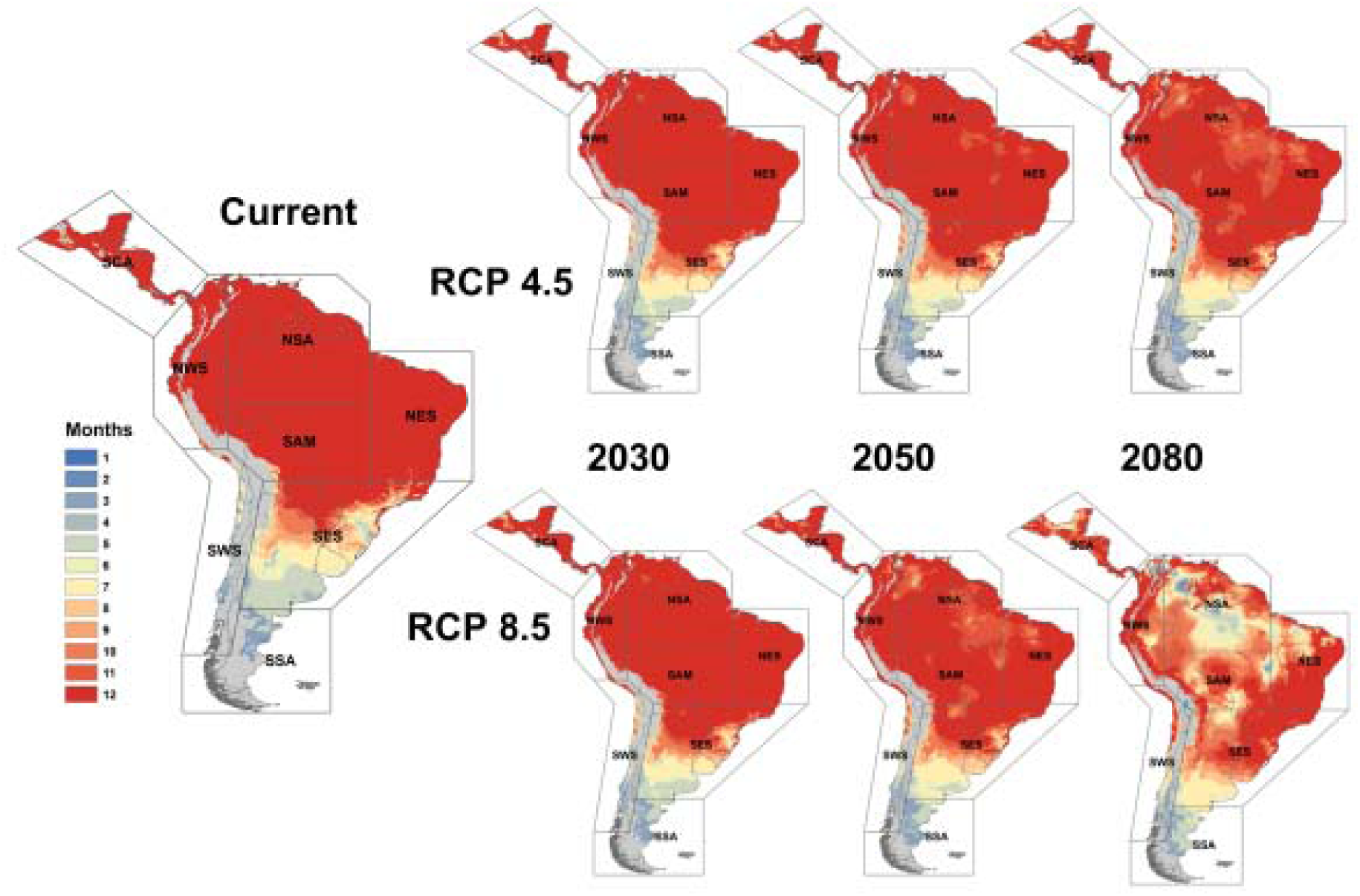
Thermal suitability for transmission of *Plasmodium vivax* malaria by the potential invasive.

**Fig. 8.**
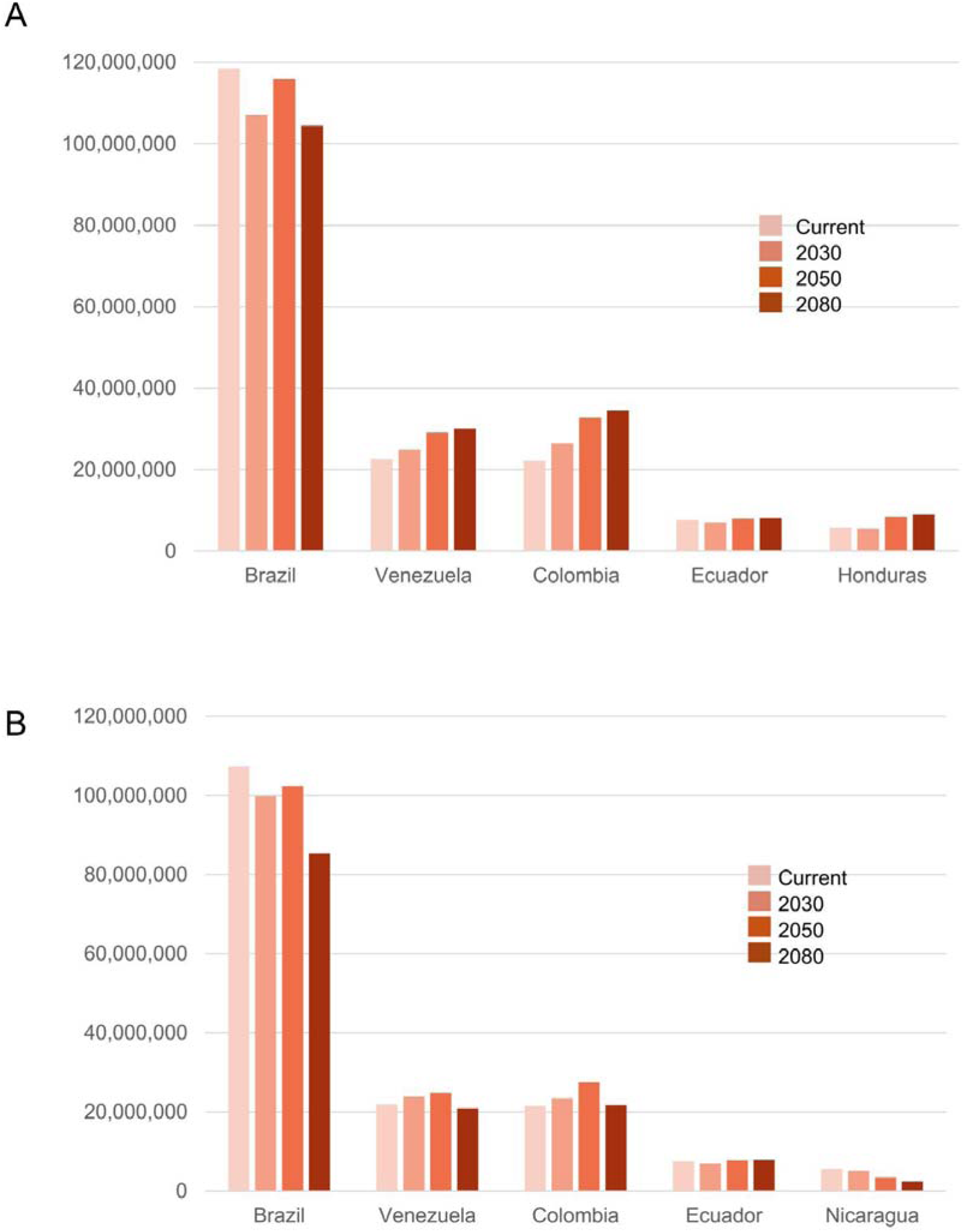
Top five countries in Central and South America with the highest population at risk (PAR) for exposure to year-round *P. falciparum* malaria (A) and *P. vivax* malaria (B) transmitted by the potential invasive *An. stephensi.* PAR will change in the future as geographic suitability shifts under climate change (RCP 4.5 shown).

### Plasmodium vivax transmitted by Anopheles stephensi

The estimated range of transmission suitability for *P. vivax* transmitted by *An. stephensi* is similar to that of *P. falciparum* transmission under current climate conditions (Fig. 7). Much of the SCA, NWS, NSA, NES, and SAM regions are suitable for year-round malaria transmission, as are northern portions of the SES region. In contrast with *P. falciparum*, the length of the *P. vivax* transmission season begins to shorten throughout the SCA, NSA, NES, and SAM regions by 2050, with the most dramatic reductions in season length projected in 2080 under RCP 8.5. Despite these reductions, modest increases in transmission suitability are projected in the southern portion of the SES region, and northern portion of the SSA region, extending into southern Argentina. Although not as high as *P. falciparium* risk, the baseline PAR for *P. vivax* transmission still exceeds that of dengue and Zika, with over 203 million PAR of year-round transmission, and over 437 million PAR for one or more months (Table 2). Although we expect some reductions in PAR under RCP 4.5 (-31.7% in SSA by 2030) and RCP 8.5 (-33.1% in SSA by 2030 under RCP), we generally expect to see steady increases in PAR for most regions, ranging from 2.0% (NES by 2030 at RCP 4.5) to 38.2% (SCA by 2080 under RCP 4.5) and 39.5% (NWS by 2080 under RCP 4.5) (Fig. 2). At the national level, an overall decrease in PAR is expected in Brazil, though Venezuela and Colombia will experience slight gains in PAR by 2050 (Fig. 7B).

### Anopheles stephensi

## Discussion

This study documents the current and projected potential exposure risk for five container breeding mosquito-pathogen pairings, encompassing three mosquito species (*Aedes aegypti, Ae albopictus, Anopheles stephensi*), and four pathogens (DENV, ZIKV, *Plasmodium falciparum*, and *Plasmodium vivax*) in Latin America. These findings describe risk for countries of Central and South America captured in the IPCC AR6 Working Group II report chapter of the same name, which describes the present-day and future thermal suitability risk of DENV and ZIKV transmission in the region [47]. This study extends and expands upon the descriptions in Castellanos et al [47], adding the potential risk from two malaria transmission scenarios in the event of invasion and introduction of the novel *Anopheles stephensi* vector. This study also disaggregates all scenarios to the country level, to allow more comprehensive examinations of baseline and future projections in the region.

We show here that a combination of projected human population changes and climate change scenarios drive vastly different responses in the number of people exposed to mosquito-borne disease transmission risk at different time horizons. These patterns depend on the underlying physiology and temperature responses of mosquito-pathogen pairings. In an increasingly urbanizing world, the capacity of mosquitoes to use artificially generated breeding habitat - containers for water storage, puddling on non-porous surfaces, ditches, garbage able to capture even small volumes of standing water - is amplified, and complicates the effects of seasonal precipitation. Thus, understanding risk as the intersection of thermal transmission suitability and human population characteristics provides us with a means to - even at a broad scale - anticipate and target vector control activity. In addition, this provides evidentiary support to develop health sector responses to climate change - e.g. to include these considerations in Health National Adaptation Plans (HNAPs). As an illustration, even when the geographic range of thermal suitability appears to be fairly consistent across most scenarios, as with dengue transmitted by *Ae. aegypti*, or *P. falciparum* malaria spread by *An. stephensi*, we still found dramatic changes in both the magnitude and geographic distribution of people at risk (PAR). As the socioeconomic pathway projections (SSPs) include spatially implicit population shifts in response to climate, these changes we found will be additionally driven by the movement of people in response to climate change [41,48].

Dengue virus transmitted by *Ae. aegypti* currently imposes major public health burdens throughout Central and South America, as evidenced by historically large outbreaks in 2019 and 2023 [49]. As shown here and elsewhere, dengue will continue to be a public health threat in the future as overall transmission suitability increases and shifts, placing new populations at risk of disease. In contrast, *Ae. albopictus*, though a competent dengue vector, is currently not as widely distributed throughout the Americas [50], but shows patterns of ongoing invasion and establishment. If this species were to become broadly established, due to its lower tolerance for high temperatures, and preference for more temperate habitats, it is currently predicted to experience greatly diminished areas of suitability in the future, corresponding to massive reductions in PAR of dengue exposure from this species. Nonetheless, in the absence of *Ae. aegypti,* it is probable that *Ae. albopictus* would become the primary vector of arboviruses [51,52]. Thus, *Ae. albopictus* presents a persistent risk for dengue transmission despite fragmented future suitability, particularly in cooler climates that would otherwise suppress transmission from *Ae. aegypti*, such as high elevations in mountainous regions [53], and currently temperate areas. Emerging precision and targeted vector control methods are becoming more prevalent and tailored to specific species, such as *Ae. aegypti* (e.g. improved sterile insect techniques [54,55], gene drive approaches [56,57], and transgenics [58])., However, these findings highlight the importance of remaining vigilant to non-target species such as *Ae. albopictus*.

*Anopheles stephensi* presents the spectre of a novel potential invader and malaria vector in the Americas. This mosquito has left its native range of the Indian subcontinent and parts of the Middle East. It has become established in urban and periurban settings across the African continent, where it has caused unprecedented urban malaria outbreaks in its introduced range [34]. In light of this global public health threat, the World Health Organization has released vector alerts concerning the spread of *An. stephensi* in 2019 [59] and in 2023 [60], underscoring the importance of preparing vector control agencies for the potential arrival of this new malaria vector.

The potential invasion of this vector is a threat to malaria elimination efforts throughout much of Latin America. *Anopheles stephensi* is a competent vector for *P. falciparum* and *P. vivax* malaria. Much of Latin America already experiences suitable temperatures for the vector and the region is projected to experience additional months of suitability and increases in PAR. Introductions and establishments in novel African locations have been attributed to possible port introductions, as well as possible land transportation route paths for goods [7]. This is equally feasible in Latin America via connections to the global trade network and multiple international ports. Clinical interventions and surveillance programs for malaria have been scaled back in some areas following elimination due to a myriad of social and ecological factors, including drug resistance, climate, agricultural expansion, waning political will, and declines in external funding [29]. However, as an urban container breeding mosquito, the control of *An. stephensi* would in many ways align with existing vector control operations for *Aedes spp.* mosquitoes, requiring minimal additional investments for capacity building and resource allocation [34]. Thus, strengthening surveillance efforts for novel *Anopheles spp* mosquitoes, added training in entomological identification, and clear communication for reporting findings of *An. stephensi*, introduced or newly established, will be essential to preparing for this potential novel vector.

To facilitate the utility of the mapping approach adopted in this study, it is essential that model outputs are available. For this work, we strive to present summary information within this study to describe the broad findings and evidence supporting the IPCC report and beyond, to extend this work to a novel potential malaria threat in a changing world. All model output is available on a public platform (XXX URL when published on Harvard Dataverse), generated at regional and country levels for all scenarios described. While we aim to add to the toolkit to confront the challenges of a changing world, communicating via maps, we hope that model output re-use can contribute to decision-making processes in other ways.

## Conclusions

The combined impacts of global change present major challenges to public health, including climate change, rapid population shifts, urbanization, and globalized movements of people, pathogens, goods, and vectors. Quantitative estimates of shifting disease transmission risks are urgently needed to target climate change adaptation planning in the health sector. In this study, we present a framework for understanding the potential shifting risk for mosquito borne diseases transmitted by container breeders in Latin America, for both currently present vectors and pathogens, and a potential invader, *Anopheles stephensi*. There is an overall trend of increased risk and geographic shifts into novel spaces. Investment in existing best practices in vector surveillance and control strategies for container-breeders, and novel control strategies will be needed to support adaptation actions and protect human health.

## Data Availability

All population product data used within are freely available online, all climate layers comprising the ensemble are freely available online, as described in methods. Please contact S.J. Ryan for gridded ensemble climate layers used. R code is available from S.J. Ryan and is a compilation of previously published code used in published articles cited within. All model outputs described within are posted on Harvard Dataverse at: (XXX URL when published).

